# Bilaterally suppressed EEG amplitudes predict death and poor functional outcomes in critically ill children

**DOI:** 10.1101/2023.11.24.23298988

**Authors:** Luisa Paul, Sandra Greve, Johanna Hegemann, Sonja Gienger, Verena Löffelhardt, Adela Della Marina, Ursula Felderhoff-Müser, Christian Dohna-Schwake, Nora Bruns

**Affiliations:** Department of Pediatrics I, Neonatology, Pediatric Intensive Care Medicine, Pediatric Neurology, and Pediatric Infectious Diseases, University Hospital Essen, University of Duisburg-Essen, Essen, Germany; C-TNBS, Centre for Translational Neuro- and Behavioural Sciences, University Hospital Essen, University of Duisburg-Essen, Essen, Germany; Department of Pediatric Cardiology/Congenital Cardiology, Heidelberg University Medical Center, Im Neuenheimer Feld 430, 69120 Heidelberg, Germany

## Abstract

**Background and objectives:** Continuous full-channel EEG is the gold standard for electrocortical activity assessment in critically ill children, but its implementation faces challenges, leading to a growing use of amplitude-integrated EEG (aEEG). While suppressed aEEG amplitudes have been linked to adverse outcomes in preterm infants and adults after cardiac arrest, evidence for critically ill children remains limited. This retrospective study aimed to evaluate the association between suppressed aEEG amplitudes in critically ill children and death or poor functional neurological outcomes.

**Methods:** 235 EEGs derived from individual patients < 18 years in the pediatric intensive care unit (PICU) at the University Hospital Essen (Germany) between 04/2014 and 07/2021 were retrospectively converted into aEEGs and amplitudes analyzed with respect to previously defined age-specific percentiles. Adjusted odds ratios for death and poor functional outcome at hospital discharge in patients with bilateral upper or lower amplitude suppression below the 10^th^ percentile were calculated accounting for neurological injuries, acute disease severity, sedation levels, and functional neurological status before acute critical illness.

**Results:** The median time from neurological insult to EEG recording was 2 days. PICU admission occurred due to neurological reasons in 43 % and patients had high overall disease severity. Thirty-three (14 %) patients died and 68 (29 %) had poor outcomes. Amplitude depression below the 10^th^ percentile was frequent (upper amplitude: 27 %, lower amplitude: 34 %) with suppression of only one amplitude less frequent than bilateral suppression. Multivariable regression analyses yielded odds between 6.63 and 15.22 for death, neurological death, and poor neurological outcomes if both upper or both lower amplitudes were suppressed. Model discrimination was excellent with areas under the curve above 0.92 for all models.

**Discussion:** This study found a high prevalence of suppressed aEEG amplitudes in critically ill children early after PICU admission, with suppression being highly associated with death and poor functional outcomes at hospital discharge. These findings emphasize the potential of early identification of high-risk PICU patients through aEEG monitoring if conventional EEG is unavailable, potentially guiding neuroprotective therapies and early neurorehabilitation.

## Introduction

Acute neurological illness accounts for up to 16 % of pediatric intensive care unit (PICU) admissions [1, 2] and an unknown proportion of PICU patients is at risk of secondary neurologic deterioration with subsequent neurologic sequelae. Evidence-based serial or continuous assessment of neurological function in critically ill children is indispensable to direct neuroprotective interventions, avoid or reduce neurologic sequelae, and preserve the developmental potential of each patient [3].

The gold standard to assess electrocortical activity is continuous full-channel EEG but suffers from high barriers to implementation even in high-resource settings [4]. To facilitate the detection of changes of EEG activity despite the absence of EEG-trained personnel, mathematical transformations of raw EEG curves are increasingly applied in intensive care settings [5-7]. These transformations provide time-compressed quantitative trend information on specific features, such as amplitude, frequency, symmetry or burst rate and are summarized with the term quantitative EEG (qEEG). The oldest type of qEEG is the amplitude-integrated EEG (aEEG), which uses a reduced set of electrodes to display the height of amplitudes [8].

Suppressed aEEG amplitudes are associated with adverse outcomes in preterm infants, asphyxiated full-term neonates, and adults after cardiac arrest [9-12]. In children, however, the general level of evidence regarding aEEG use is still low [13]. Only recently, reference values for healthy children have been published [14, 15] and there is no more but incipient evidence from small studies that amplitude suppression may be associated with death or adverse outcomes as shown in neonates and adults [16-18]. Yet, aEEG devices are ubiquitous available in pediatric departments due to their indispensable role for the diagnostic and therapeutic decision-making path in neonatal asphyxia. Pediatric intensive care teams become increasingly familiar with aEEG devices, promoting this type of EEG monitoring [19, 20].

The aim of this study was to assess the association of suppressed aEEG amplitudes in critically ill children with death and functional neurological short-term outcome. We retrospectively processed aEEGs from conventional EEGs that were conducted in our PICU and investigated the height of amplitudes as a predictor for death or poor functional shortterm outcome.

## Methods

### Study setting and eligibility

Retrospective observational study using EEGs conducted in the PICU of the University Hospital Essen (Germany). The children’s hospital EEG database was screened for EEGs conducted in the PICU since its implementation in April 2014 until July 2021. All EEGs conducted in patients < 18 years in the PICU during the study period were assessed for eligibility. If a patient received several EEGs during the same PICU stay, only the first EEG was eligible for analysis.

### Clinical data and outcomes

Clinical data were retrieved by review of digitalized patient charts and from the electronic patient documentation system.

- Acute disease severity at PICU admission was quantified using the pediatric sequential organ failure assessment (pSOFA) [21]. Patients with pSOFA scores above 2 are considered at high risk for mortality.
- Acute neurological injuries were identified from cranial imaging findings and categorized into generalized and focal lesions (Table 1).
- Sedation levels were categorized based on the substance, dosage, and type of administration (continuous versus intermittent) (Table 2).
- Functional neurological status before acute critical illness and at hospital discharge was determined by the pediatric cerebral performance category (PCPC) [22]. Neurological outcome was categorized as good (PCPC 1 – 3 = no, mild or moderate disability), poor (PCPC 4 – 6 = severe disability, persistent vegetative state or brain death/death), and deceased (subset of the poor outcome group). According to the original publication [22], a PCPC score of 6 is assigned only for brain death. However, formal brain death diagnostics are not routinely carried out in our center unless postmortal organ donation is a potential scenario or relatives are reluctant to withdraw life-sustaining therapies in causes with poor prognosis. Thus, death from any reason was assigned a score of 6 on the PCPC. Only in-hospital death during the same hospital stay was considered as death for this study. Neurological death was defined as death from cerebral injury (primary or secondary), in contrast to, e.g., multi-organ failure.

**Table 1:** Sedation grading.

| Grade | Administered medication |
| --- | --- |
| 0 | No sedatives |
| 1 | SD only (benzodiazepine, propofol, ketamine, chloral hydrate) |
| 2 | Continuous infusion of propofol ± SD |
| 3 | Continuous infusion of opiate |
| 4 | Continuous infusion of opiate + benzodiazepine ± SD |
| 5 | Continuous infusion of opiate + benzodiazepine + katemine ± SD |
| 6 | Inhalative sevoflurane / continuous infusion of thiopental |

SD = single doses

**Table 2:**
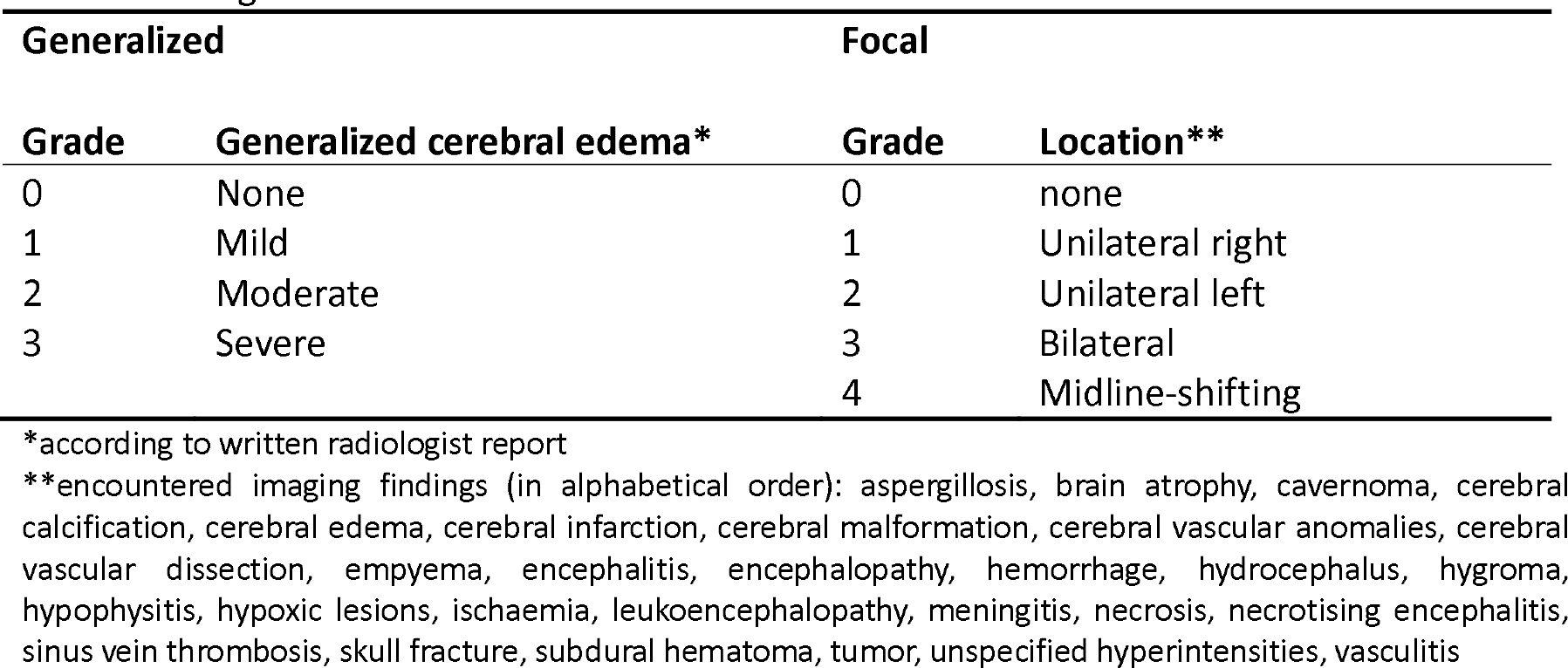
Categorization of cerebral lesions.

| Generalized |  | Focal |  |
| --- | --- | --- | --- |
| Grade | Generalized cerebral edema* | Grade | Location** |
| 0 | None | 0 | none |
| 1 | Mild | 1 | Unilateral right |
| 2 | Moderate | 2 | Unilateral left |
| 3 | Severe | 3 | Bilateral |
|  |  | 4 | Midline-shifting |

\*according to written radiologist report
\*\*encountered imaging findings (in alphabetical order): aspergillosis, brain atrophy, cavernoma, cerebral calcification, cerebral edema, cerebral infarction, cerebral malformation, cerebral vascular anomalies, cerebral vascular dissection, empyema, encephalitis, encephalopathy, hemorrhage, hydrocephalus, hygroma, hypophysitis, hypoxic lesions, ischaemia, leukoencephalopathy, meningitis, necrosis, necrotising encephalitis, sinus vein thrombosis, skull fracture, subdural hematoma, tumor, unspecified hyperintensities, vasculitis

### Local practice of EEG recording

In our PICU, full channel EEG is available only at daytime on regular working days except for acute emergencies, for which it is available at any time. During off-hours, continuous reduced channel EEG and aEEG is conducted in acute neurologically ill patients or those at risk for neurological complications. In patients with continued neurological deficits, coma, or high risk for adverse neurological outcomes, the reduced channel EEG is interrupted to enable full channel conventional recording EEG as soon as possible on the next regular working day. All EEGs analyzed for this study were recorded as full channel conventional EEGs.

### EEG conduction and aEEG conversion

Full-channel EEGs were conducted according to the international 10–20 system after skin preparation with OneStep EEG Gel Abrasiv plus®. Impedance was checked and skin preparation repeated until impedances were <5 kΩ for all electrodes, as required by the German Society for Clinical Neurophysiology and Functional Imaging. All EEGs were recorded using Neurofax EEG devices and polaris.one software v4.0.4.0 (Nihon Kohden, Tokyo, Japan). aEEGs were computed with Polaris Trend Software QP-160AK (Nihon Kohden, Tokyo, Japan).

### Amplitude assessment

The C3-P3 and C4-P4 channels were analyzed channels semi-automatically by two independent researchers (LP and NB) as previously described in detail [14, 23]. Briefly, amplitudes were measured by aligning the integrated measurement tool with the upper and lower amplitude. Next, the mean value between the two raters was calculated and used for further calculations and analyses.

Based on previously specified age-specific percentiles [14], the upper and lower amplitudes were classified as suppressed (<10^th^ percentile) or normal (≥ 10^th^ percentile) for each hemisphere. Suppression patterns were then classified into normal (no amplitude suppressed), one amplitude suppressed, or both amplitudes suppressed for the upper and lower amplitudes, respectively.

### Statistical analyses

Descriptive analyses are presented as mean ± standard deviation (SD) for continuous normally distributed data; skewed data are presented as median and interquartile range (IQR). Discrete variables are summarized as counts and relative frequencies.

Interrater agreement was assessed by Bland-Altmann plots [24] and intraclass correlation coefficients (two-way mixed model for individual ratings (ICC 3,1)) [25] for the upper and lower amplitudes as previously described [14, 23].

Adjusted and unadjusted odds ratios with 95 % confidence intervals (CI) for death and poor functional outcome were calculated by multivariable logistic regression analyses comparing patients with no versus bilateral amplitude suppression. Adjustment sets were identified by causal diagrams based on the theory of directed acyclic graphs (DAG) [26, 27], as recommended for empirical research in pediatrics and intensive care medicine [28, 29] (Figure 1) and comprised the type of acute neurologic injury (Table 1), sedation levels at the time of recording (Table 2), acute overall disease severity (pSOFA score), and neurological functional status before the acute critical illness (PCPC).

**Figure 1:**
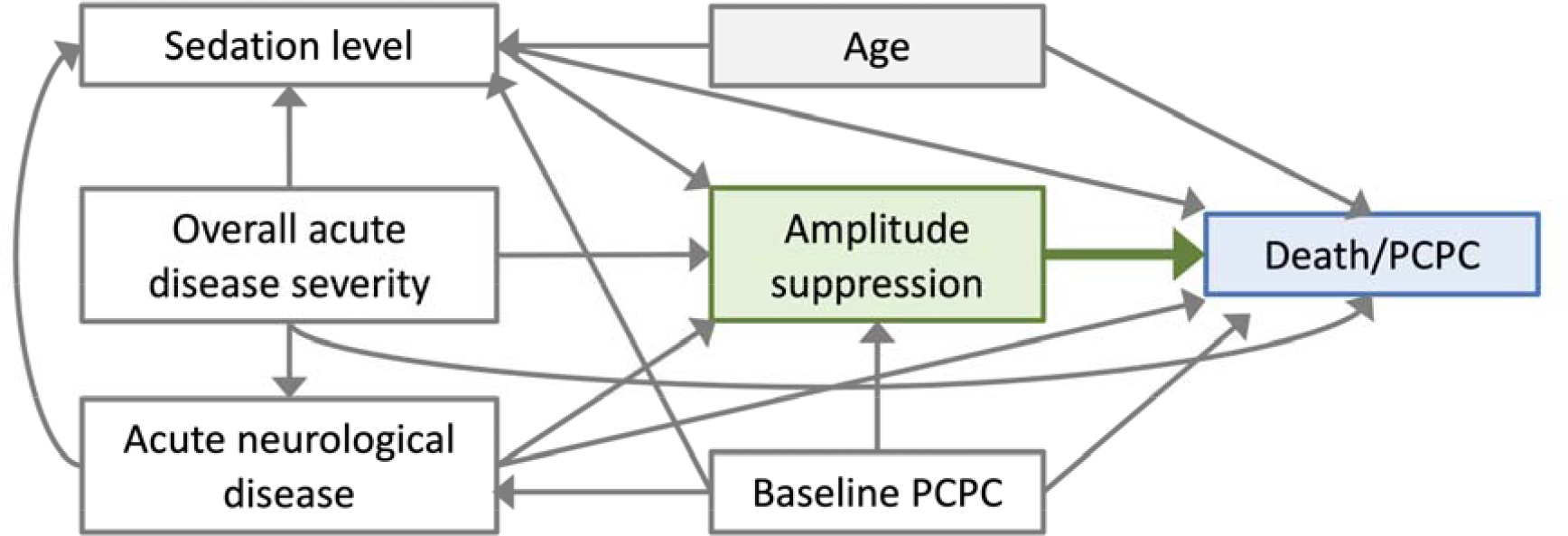
Causal diagram to identify the adjustment set for regression analyses. PCPC = Pediatric cerebral performance category. Box colors: green = exposure, blue = outcome, white = variables that must be adjusted for, grey = no adjustment necessary.

SAS Enterprise Guide 8.3 (SAS Institute Inc., Cary, NC, USA) was used for statistical analyses and figure production.

### Ethics approval

The study was approved by the local Ethics Committee of the Medical Faculty of the University of Duisburg-Essen (21-10264-BO). Written informed consent was waived according to local legislation for analysis of retrospective anonymized data.

## Results

In total, 235 EEGs from 235 individual patients with a mean duration of 16:46 min (± 6:15 min) were analyzed (Figure 2). The median time between neurological insult and PICU admission was 0 days and median time from neurological insult to EEG recording was 2 days. The included patients displayed high degrees of acute overall and neurological illness with median pSOFA scores above 2 and neurological reasons for PICU admission in 101 cases (43 %) (Table 3). Thirty-three (14 %) patients died and 68 (29 %) had poor outcomes. Patients with good outcome were older, had lower pSOFA scores, were mechanically ventilated and sedated less frequently, and had shorter durations of PICU and hospital stay. Across groups, most children had previously good neurological function with baseline PCPC scores differing only in the upper quartiles.

**Table 3:** Patient characteristics by outcome group.

|  | Outcome |  |  |
| --- | --- | --- | --- |
|  | Good<br>(n = 167) | Poor<br>(n = 68) | Deceased*<br>(n = 33) |
| Female, *n* (%) | 70 (42 %) | 26 (38 %) | 13 (39 %) |
| Age (years), *median (IQR)* | 3 (0 – 12) | 2 (0 – 7.5) | 0 (0 – 5) |
| Weight (kg), *median (IQR)* | 15.0<br>(7.0 – 42.6) | 12.0<br>(6.4 – 21.2) | 10.0<br>(6.0 – 19.4) |
| Reason for PICU admission |  |  |  |
| Acute neurologic disease | 65 (39 %) | 20 (29 %) | 6 (18 %) |
| Cerebral injury or lesion (trauma, bleeding, tumor) | 16 (10 %) | 8 (12 %) | 6 (18 %) |
| Hypoxia/resuscitation | 2 (1 %) | 6 (9 %) | 5 (15 %) |
| Organ failure (non-neurologic) | 30 (18 %) | 9 (13 %) | 7 (21 %) |
| Other | 54 (32 %) | 25 (37 %) | 9 (27 %) |
| Days between PICU admission and insult, *median (IQR)* | 0 (0 – 0) | 0 (0 – 0) | 0 (0 – 0) |
| Days insult to EEG, *median (IQR)* | 2 (1 – 5) | 2 (1 – 6) | 2 (1 – 6) |
| pSOFA score | 4 (2 – 6) | 7 (5 – 11) | 9 (6 – 11) |
| Cardiopulmonary resuscitation prior to EEG | 10 (6 %) | 17 (25 %) | 11 (33 %) |
| Invasive mechanical ventilation, *n* (%) | 33 (20 %) | 41 (60 %) | 28 (85 %) |
| Administration of sedatives within 24 h before or during recording | 48 (29 %) | 41 (61 %) | 24 (73 %) |
| Sedation by continuous infusion | 47 (28 %) | 40 (60 %) | 24 (73 %) |
| Antiepileptic drug treatment at time of EEG** | 12 (7 %) | 5 (7 %) | 2 (6 %) |
| PCPC baseline, *median (IQR)* | 1 (1 – 1) | 2 (1 – 3) | 1 (1 – 2) |
| PCPC ICU discharge, *median (IQR)* | 2 (1 – 3) | 5 (4 – 6) | 6 (6 – 6) |
| PCPC hospital discharge, *median (IQR)* | 2 (1 – 2) | 5 (4 – 6) | 6 (6 – 6) |
| PCPC decline baseline to hospital discharge, *median (IQR)* | 0 (0 – 1) | 3 (1 – 5) | 5 (4 – 5) |
| Length of PICU stay (days), *median (IQR)* | 10<br>(4 – 20) | 16<br>(5 – 28) | 9<br>(3 – 19) |
| Length of hospital stay (days), *median (IQR)* | 18<br>(9 – 34) | 25<br>(8 – 42) | 9<br>(3 – 20) |
| Death, *n* (%) | 0 (0 %) | 33 (49 %) | 33 (100 %) |
| Neurological death, *n* (%) | 0 (0 %) | 24 (35 %) | 24 (73 %) |

\* Subset of poor outcome group; \*\* In addition to sedation; IQR = interquartile range, PICU = pediatric intensive care unit, pSOFA = pediatric sequential organ failure assessment

**Figure 2:**
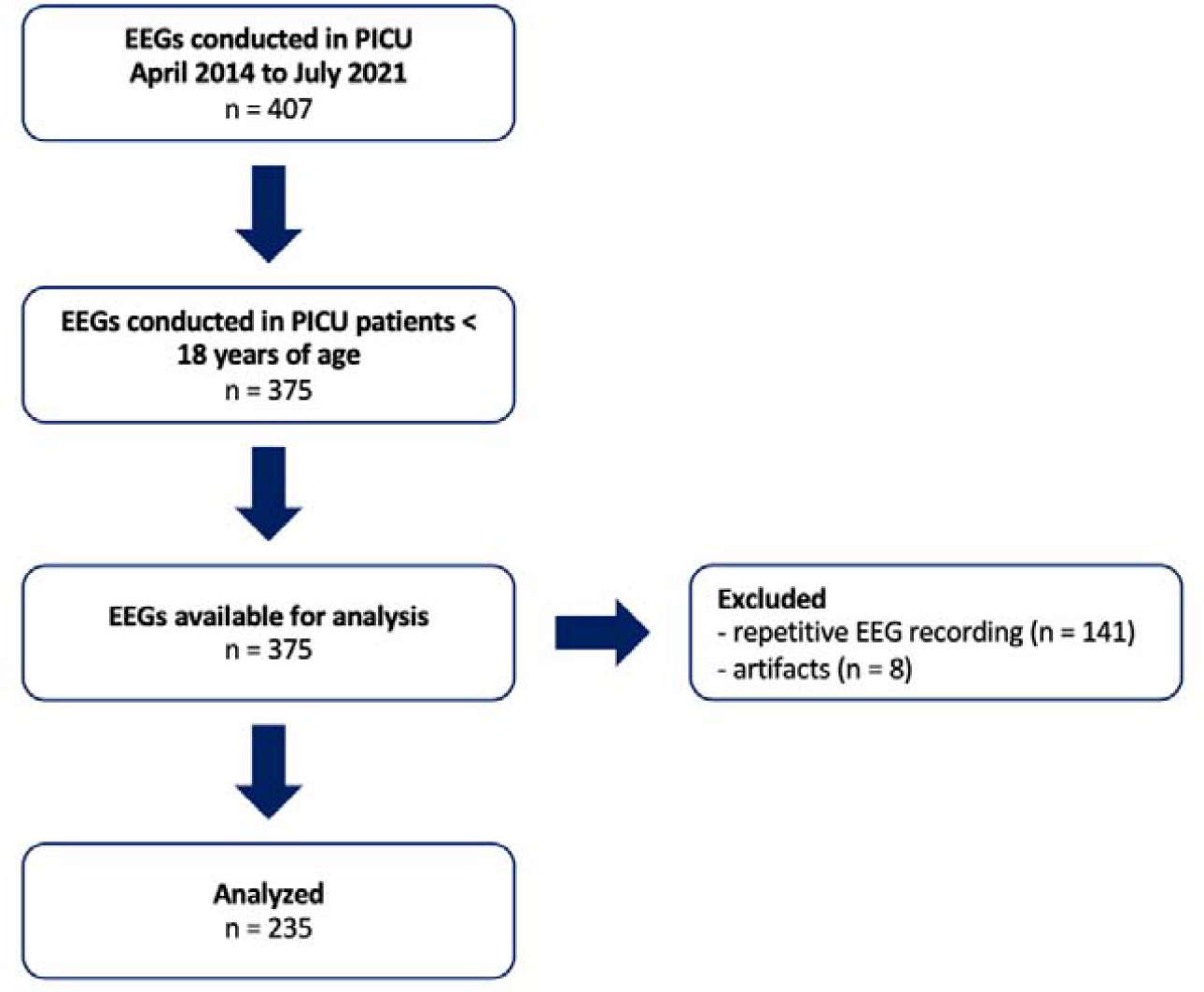
Screened and anayzed EEGs PICU = pediatric intensive care unit

The intraclass correlation coefficients were 0.94 both for the upper and lower amplitudes. The mean interrater difference for the upper amplitude was 0.2 μV (± 8.0 μV) and 0.6 μV (± 1.7 μV) for the lower amplitude.

Grouping by amplitude suppression pattern showed that amplitude depression below the 10^th^ percentile was frequent (upper amplitude: 27 %, lower amplitude: 34 %) and that suppression of only one amplitude was less frequent than bilateral suppression (Table 4). Death and poor neurological outcomes were highest if both amplitudes were suppressed and lowest in the group with no amplitude suppression (Table 4, figure 3). Multivariable regression analyses yielded odds between 6.63 and 15.22 for death, neurological death, and poor neurological outcomes if upper or lower amplitudes were suppressed (Figure 4). Model discrimination was excellent with areas under the curve above 0.92 for all models (Supplementary table S1). Unadjusted odds ratios were similarly high but with suboptimal discriminatory performance (Supplementary table S1)

**Table 4:** Association between amplitude suppression and outcome.

| Amplitude | Suppression pattern | Outcome |  |  |
| --- | --- | --- | --- | --- |
|  |  | Good | Poor | Deceased* |
| Upper | Both normal | 103 (84 %) | 20 (16 %) | 5 (4 %) |
|  | One suppressed | 27 (71 %) | 11 (29 %) | 4 (11 %) |
|  | Both suppressed | 37 (50 %) | 37 (50 %) | 24 (32 %) |
| Lower | Both normal | 87 (86 %) | 14 (14 %) | 3 (3 %) |
|  | One suppressed | 36 (86 %) | 6 (9 %) | 2 (5 %) |
|  | Both suppressed | 44 (48 %) | 48 (52 %) | 28 (30 %) |

All numbers represent n (%); \*Subset of poor outcome group

**Figure 3:**
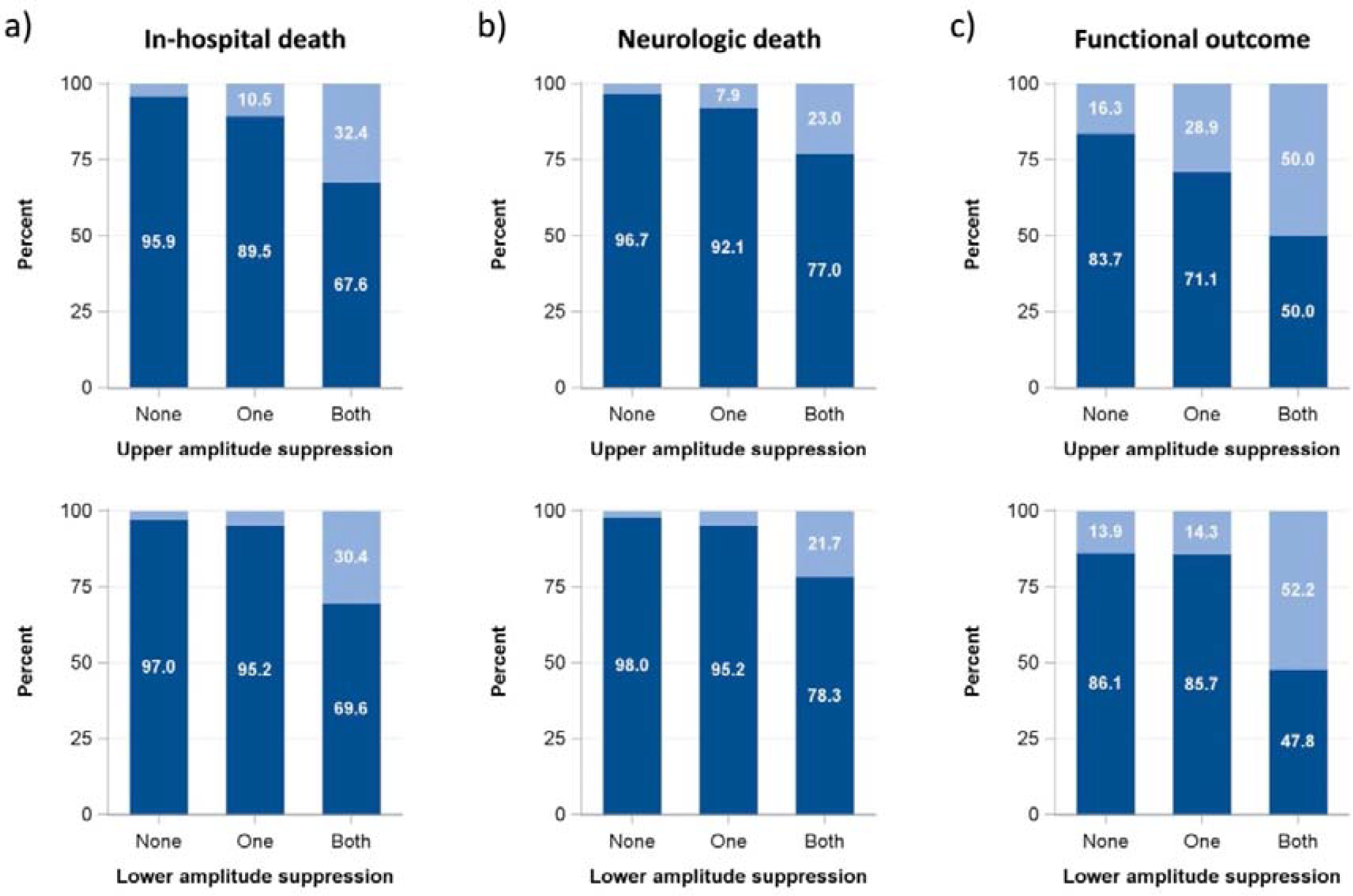
EEG amplitude suppression and outcomes in critically ill children a)In-hospital death (dark blue = survived, light blue = deceased) b)Neurological death (dark blue = no neurological death, light blue = neurological death) c)Functional outcome ((dark blue = good outcome (PCPC 1 – 3), light blue = poor outcome (PCPC 4 – 6))

**Figure 4:**
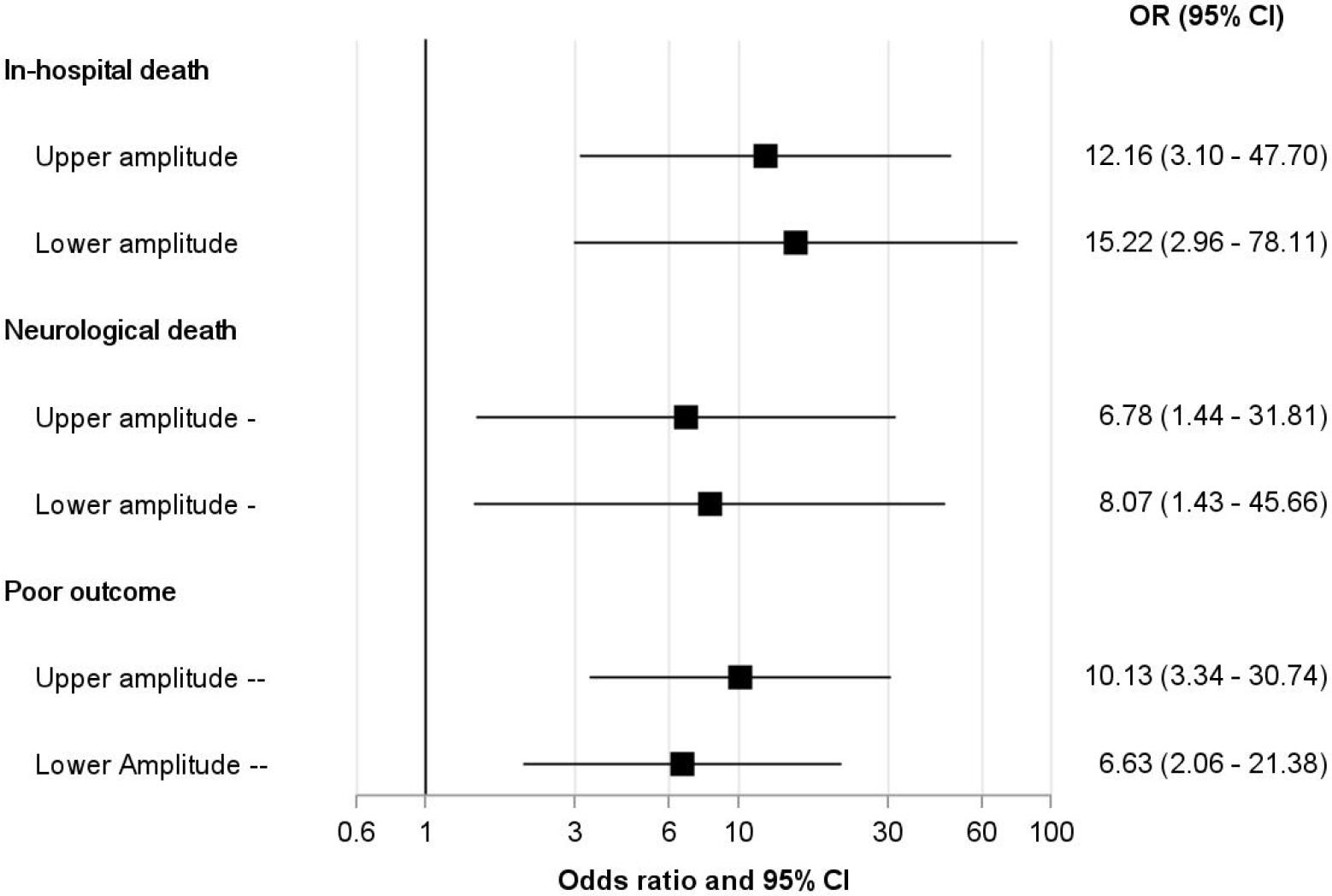
Adjusted odds ratios for death, neurological death, and poor outcome in critically children with suppressed EEG amplitudes CI = confidence interval, OR = odds ratio

## Discussion

This study found suppression of EEG amplitudes in a high proportion of critically ill children early after PICU admission. Suppressed amplitudes were associated with death and poor functional outcome at discharge in descriptive and regression analyses. Discrimination of outcomes was enhanced when baseline functional status, neurological lesions, acute disease severity, and sedation levels were accounted for. Overall, odds for death and poor functional outcome were strongly increased if both amplitudes were below the 10^th^ percentile for age. This study confirms emerging evidence from three smaller studies on the association between aEEG amplitudes and outcomes in PICU patients with meningitis and after cardiac arrest [16-18]. It further aligns with adult reports on conventional EEG which linked unfavorable outcomes to worse background patterns, unreactive or flat EEGs, and burst suppression [30, 31].

The patient population in this study was highly preselected, with high overall disease severity and additionally high burden of neurologic disease compared to average PICU patients [1, 2], putting these patients at special risk for death and neurologic sequelae. Our results highlight the potential of aEEG to monitor and identify high-risk PICU patients early during the course of disease. Early identification of these patients may indicate the need for advanced neuromonitoring and potentially warrant neuroprotective strategies and early neurorehabilitation. The differing discriminatory performance between adjusted and unadjusted regression models points out the importance of additional clinical information for aEEG interpretation. Of special importance in PICU patients are sedation levels and neuroactive medication, which affect EEG activity [32-35] and can induce severe amplitude suppression [36, 37]. Hemodynamic changes also affect electrocortical activity [13], possibly via changes of oxygen supply and metabolic clearance.

In contrast to other studies in PICU patients investigating continuous EEG monitoring and amplitudes over time, this study assessed only one short EEG per patient conducted early after PICU admission. Therefore, the predictive potential of amplitude evolution over time could not be investigated. On the other hand, this study shows that accompanying conditions are extremely important to account for. In the acute phase of critical illness, the interplay of vasopressor/inotrope administration, sedation levels, and organ dysfunction is highly dynamic, warranting advanced time series approaches to accurately account for changes of accompanying physiological and treatment conditions in long-term recordings. The “static” approach of this study made a large-scale aEEG assessment in very severely ill children possible for the first time, thereby providing important evidence for clinicians’ daily practice. Another limitation of this study is the fact that by using the 10^th^ percentile as a threshold to define amplitude suppression, 10 % of healthy children physiologically display amplitudes that were classified as suppressed in this study. Compared to a lower percentile as a threshold, this reduces specificity in favor of sensitivity. Larger scale studies should identify optimum thresholds to define amplitude suppression, either by using age-specific percentiles or absolute values. To optimize prediction and discrimination of death and functional outcome by EEG, further quantitative EEG parameters or a combination of several parameters should also be systematically investigated in this context. Due to the retrospective character of this study, the functional outcome assessment was limited to the PCPC as a simple and common measure of PICU outcomes. Prospective investigations should also assess cognitive, emotional, and social health functioning, as their impairment contributes to post-PICU sequelae, such as the post-intensive care syndrome [38-40].

The most important finding of this study is that early aEEG suppression is strongly associated with adverse outcome in critically ill children and that aEEG interpretations needs to account for neurological lesions, acute disease severity and neuroactive medication. Further research is needed to investigate even earlier aEEG recordings and the evolution of amplitudes over the course of disease. Further qEEG parameters should be examined regarding their potential improve identification of high-risk patients and direct neuroprotective therapeutic and neuro-rehabilitative strategies.

## Data availability

Anonymized data not published within this article will be made available by request from any qualified investigator.

---

**Supplementary table S1:** Adjusted and unadjusted odds ratios to predict outcomes and discriminatory model performance.

| Outcome | Amplitude | Adjusted | AUC | HL test (p) | Unadjusted | AUC | N included in adjusted/unadjusted model |
| --- | --- | --- | --- | --- | --- | --- | --- |
|  |  | Odds ratio (95 % CI) |  |  | Odds ratio (95 % CI) |  |  |
| In-hospital death | Upper | 12.16<br>(3.10 – 47.70) | 0.94 | 0.86 | 11.33<br>(4.09 – 31.37) | 0.77 | 192/197 |
|  | Lower | 15.22<br>(2.96 – 78.11) | 0.92 | 0.99 | 14.29<br>(4.17 – 48.95) | 0.75 | 186/193 |
| Neurological death | Upper | 6.78<br>(1.44 – 31.81) | 0.95 | 0.97 | 8.87<br>(2.86 – 27.58) | 0.74 | 192/197 |
|  | Lower | 8.07<br>(1.43 – 45.66) | 0.93 | 0.99 | 13.75<br>(3.12 – 60.70) | 0.74 | 186/193 |
| Poor outcome | Upper | 10.13<br>(3.34 – 30.74) | 0.93 | 0.90 | 5.15<br>(2.66 – 9.98) | 0.69 | 192/197 |
|  | Lower | 6.63<br>(2.06 – 21.38) | 0.93 | 0.93 | 6.78<br>(3.38 – 13.61) | 0.72 | 186/193 |

